# Clinical and Decisional Trajectories Preceding Early Postprocedural Death Among Patients with Preexisting Code-Status Limitations

**DOI:** 10.64898/2026.09.11.26362839

**Authors:** Matthew B. Allen, Jocelyn L. Streid, Elizabeth J. Lilley, Christine S. Ritchie, Christy E. Cauley, Jason N. Batten, Rachelle E. Bernacki, Amanda J. Reich, Preeti R. John, David L. Hepner, Angela M. Bader, Sachin J. Shah

## Abstract

**Objective:** To develop a typology of recurrent clinical and decisional trajectories preceding early postprocedural death among patients with preexisting code-status limitations.

**Background:** Patients with do-not-resuscitate orders experience higher postoperative mortality, but the clinical and decisional processes underlying early deaths are poorly understood. Mortality and code status alone cannot determine whether these deaths reflect constrained rescue, unsuccessful treatment, progression of terminal illness, or reassessment-driven redirection of care.

**Methods:** We conducted a retrospective qualitative trajectory analysis of electronic health record documentation across five academic and community hospitals from March 2024 through June 2025. Eligible adults had an active preprocedural code-status limitation and died within 3 calendar days of a procedure requiring anesthesia, before hospital discharge. Using an adapted sequence-based composite approach, 2 reviewers independently reconstructed cases as ordered clinical and decisional sequences and reached consensus on recurrent trajectory families; illustrative composites were then generated. A third reviewer independently classified cases while blinded to the consensus assignments; agreement was assessed using unweighted Cohen κ.

**Results:** Among 2,833 patients with preexisting code-status limitations who underwent a procedure, 44 (1.6%) died within 3 days; the 42 who died before hospital discharge were included. Four trajectory families were identified: established terminal trajectory (5 [12%]), abrupt terminal event (5 [12%]), comfort-focused redirection after reassessment (29 [69%]), and sustained rescue attempt (3 [7%]). Redirection followed progressive deterioration or persistent critical illness despite treatment in 20 cases and a discrete event or finding in 9; rescue-oriented treatment frequently continued or escalated before reassessment and redirection. In sustained-rescue trajectories, intensive treatment continued through death, including in 2 patients whose limitations on CPR remained in effect. The independent reviewer agreed with the consensus classification for 39 of 42 cases (92.9%; κ, 0.86; 95% CI, 0.70–1.00).

**Conclusions:** Early postprocedural deaths among patients with preexisting code-status limitations followed distinct clinical and decisional trajectories. Mortality and code status alone could not distinguish an established terminal trajectory, abrupt death, reassessment-driven redirection, or sustained but unsuccessful rescue. A longitudinal approach that considers procedural intent, evolving prognosis, treatment, and reassessment provides a more clinically meaningful basis for interpreting mortality and structuring perioperative decision making.

**MINI ABSTRACT:** Among patients with preexisting code-status limitations who died within 3 days of a procedure, four recurrent trajectories differed in relationships among baseline illness, clinical developments, prognostic reassessment, rescue-oriented treatment, and changes in treatment focus. These longitudinal dynamics were essential to interpreting early postprocedural death.

## INTRODUCTION

Patients with do-not-resuscitate (DNR) orders consistently experience higher postoperative mortality than patients without such orders.^1–3^ For surgeons and anesthesiologists, this association raises a practical and ethical concern: that continuing treatment limitations around the time of surgery may allow patients to die by limiting the scope of rescue after potentially reversible complications.^4,5^ Yet this association may also reflect greater underlying illness burden^6^ or decisions to redirect treatment after events prompt reassessment of prognosis, treatment burden, and goals of care. Distinguishing among these mechanisms is essential because they imply different interpretations of mortality and different targets for perioperative decision making and quality improvement.

Prior registry studies could not reliably differentiate these explanations because they classified DNR status as a fixed preoperative exposure and lacked temporally detailed data on perioperative code-status decisions and subsequent transitions in the focus of care.^1–4^ Our prior multicenter quantitative analysis narrowed this uncertainty by comparing patients who all presented for a procedure with preexisting code-status limitations but differed in whether those limitations were maintained or temporarily reversed.^7^ We found that many early postprocedural deaths followed a transition to comfort-focused care, indicating that excess mortality should not be interpreted exclusively as evidence of missed opportunities for rescue.^7^ However, code-status transitions in isolation do not reveal the clinical events, circumstances, and deliberations that produced them—information needed to interpret perioperative mortality in relation to high-stakes treatment decisions.

To address this gap, we conducted a qualitative trajectory analysis of hospitalized patients with preexisting code-status limitations who died within 3 days after a procedure. We sought to develop a typology of recurrent clinical and decisional trajectories preceding early postprocedural death and to characterize how prognosis, procedural intent, clinical developments, and treatment decisions unfolded within each trajectory.

## METHODS

### Design, Setting, Participants

We conducted a retrospective qualitative trajectory analysis of electronic health record (EHR) documentation to reconstruct the clinical and decisional courses of adults with preexisting code-status limitations who received anesthesia care for a procedure at five academic and community hospitals within Mass General Brigham from March 2024 through June 2025.

We defined a preexisting code-status limitation as an active preprocedural order limiting CPR, with or without an additional limitation on intubation. We focused on patients with preexisting code-status limitations because the American College of Surgeons (ACS) and American Society of Anesthesiologists recommend explicit reconsideration of existing resuscitation directives in the perioperative period.^8,9^

Patients remained eligible whether the preexisting limitation was maintained, modified, or temporarily reversed for the procedure. We included patients who remained hospitalized and died within 3 calendar days; patients who died after routine discharge were excluded because their terminal trajectory could not be reconstructed. Sample size was determined by the finite cohort rather than thematic saturation.

We focused on deaths within 3 days to preserve temporal proximity among the procedure, early postprocedural course, and treatment decisions. Later deaths may involve prolonged critical illness and repeated reassessment requiring a distinct analytic frame. The Mass General Brigham Institutional Review Board determined that the study met criteria for exemption from review. We report this study in accordance with the Standards for Reporting Qualitative Research (SRQR).^10^

### Data Sources and Chart Review

We identified eligible patients from the previously described cohort derived from the Mass General Brigham Patient Data Registry, a repository of clinical and administrative data.^7^ The source cohort included adults undergoing a procedure requiring anesthesia who had a non–full code status at the preoperative evaluation on the day of the procedure. Hospitalization and death within 3 days were identified from structured registry data and confirmed through manual record review. We reviewed records from hospital admission or, for patients not already hospitalized, the preoperative anesthesia evaluation on the day of the procedure, through death. Reviewed sources included anesthesia and procedural records, clinical notes, goals-of-care and family-meeting documentation, and death summaries. Before pilot abstraction, MA and JS specified six domains reflecting successive phases of the perioperative course: baseline health status, procedural context, preprocedural decision making, intraprocedural course, postprocedural course and decision making, and terminal outcome (**Figure 1**). Direct identifiers were excluded from materials used for cross-case comparison and composite construction.

**Figure 1.**
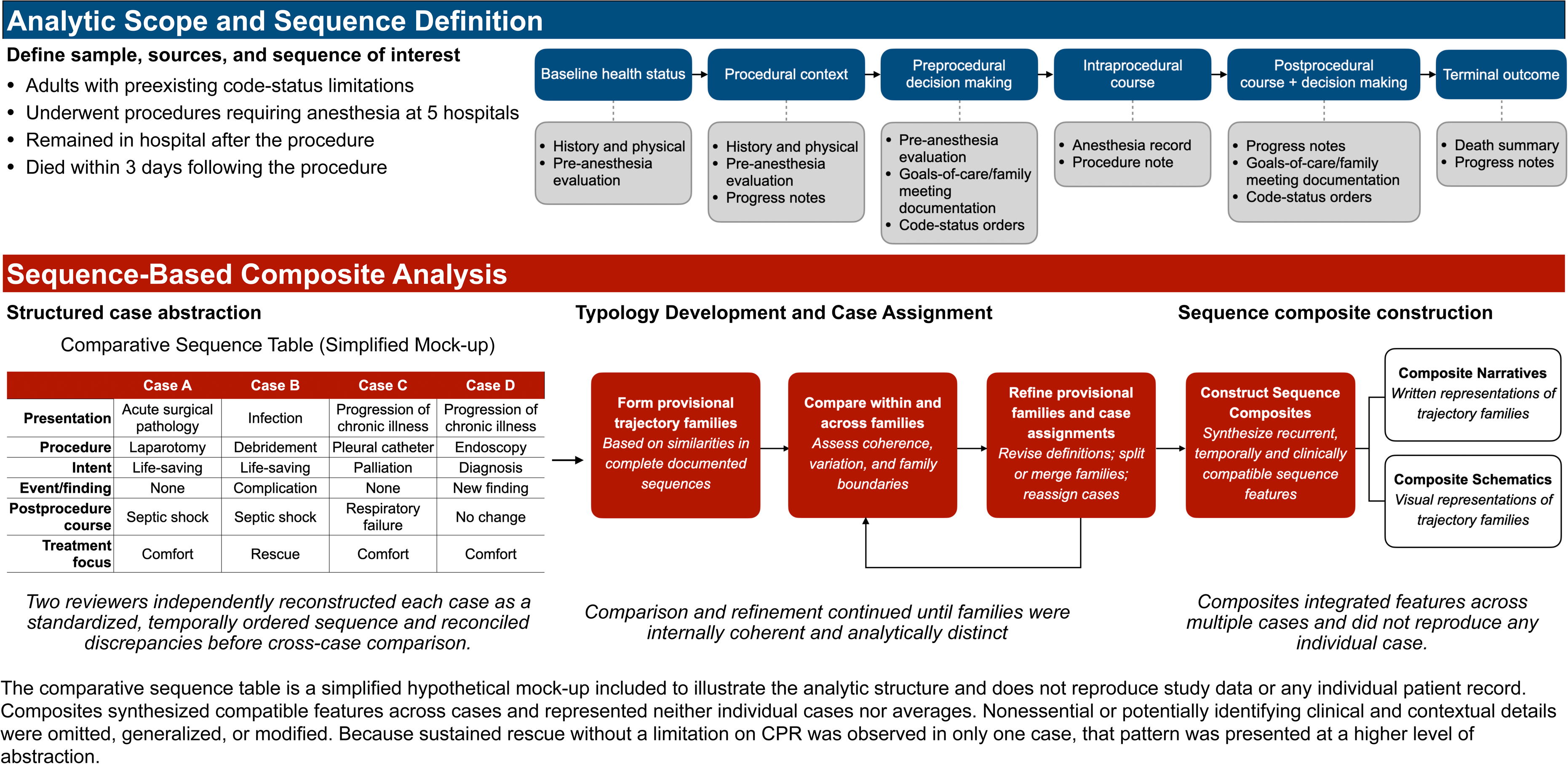
Methodological Overview of Sequence-Based Composite Analysis.

### Qualitative Trajectory Analysis Using Sequence-Based Composites

We used an adapted sequence-based composite (SBC) approach, a case-oriented qualitative method representing each case as an ordered clinical and decisional sequence.^11^ The primary output was a typology of recurrent trajectory families, each illustrated by a composite narrative and schematic synthesizing temporally compatible features rather than an individual case or average.

### Structured Case Abstraction

MA and JS independently reviewed 6 randomly selected cases to develop the abstraction guide and codebook within the six prespecified domains. During pilot review, they inductively added and refined codes for recurrent clinical events and findings, decisions, and potential turning points. They reconciled differences and incorporated study-team feedback before full-cohort abstraction.

Using the revised guide and codebook, MA and JS independently reconstructed each case as a temporally ordered sequence of clinical events and findings, evolving prognostic understanding, treatments, decisions, changes in treatment focus, and death. They distinguished discrete events from progressive deterioration or nonresponse and identified potential turning points involving changes in prognosis, available treatment options, or goals of care. Reassessment required documented reconsideration of prognosis, treatment options, or goals; transition to comfort-focused care required documentation of a broader shift in treatment focus. Features not supported by documentation were coded as ambiguous rather than inferred. MA and JS reconciled discrepancies through review of the source record and entered the agreed sequences into a standardized table.

### Typology Development and Case Assignment

MA and JS independently assigned cases to provisional families based on similarities across complete documented sequences; diagnosis, procedure type, code status, and terminal outcome informed but did not determine assignment. Through iterative within- and between-family comparison, they reconciled assignments, refined definitions, identified within-family variation, and split or merged families or reassigned cases as needed for internal coherence and analytic distinction. They reached consensus on final definitions and assignments and maintained an audit trail of revisions, boundary cases, and rationales.

### Sequence Composite Construction

After finalizing the trajectory families, MA synthesized recurrent sequence features into an illustrative narrative and schematic for each family, including only features that were temporally and clinically compatible across cases. Each narrative summarized the family’s clinical and decisional sequence, while the corresponding schematic represented qualitative changes in prognostic certainty and the relative order of clinical developments, decisions, and death.

Potentially identifying clinical and contextual details were omitted, generalized, or modified. Because sustained rescue without a limitation on CPR was observed in only one case, that pattern was presented at a higher level of abstraction. MA revised the composites after multidisciplinary team members with expertise in surgery, medicine, anesthesiology, geriatrics, and palliative care (EL, CC, SS, JS, and RB) reviewed them for clinical coherence and plausibility.

### Reproducibility Assessment

After the family definitions and consensus case assignments were finalized, JB—an anesthesiologist and critical care physician who had not participated in case abstraction, typology development, or composite construction—independently assigned each standardized sequence, presented in randomized order, to one trajectory family. JB was blinded to the consensus assignments and received the family definitions, including defining and associated features, together with the composite narratives and schematics.

We compared JB’s assignments with the MA–JS consensus assignments using unweighted Cohen κ with a 95% confidence interval and overall percentage agreement. Analyses were conducted at the family level, with subtypes grouped within their parent family. After calculating agreement, MA and JB reviewed discrepant cases to identify reasons for disagreement and determine whether the family definitions or composite presentations required clarification. This review did not alter the assignments used to calculate agreement.

## RESULTS

Among 2,833 patients with preexisting code-status limitations who underwent a procedure during the study period, 44 (1.6%) died within 3 days.^7^ Of these, 42 died before hospital discharge and were included; 2 died at home after routine discharge and were excluded because their terminal trajectories could not be reconstructed. Patient characteristics and clinical context are summarized in **Table 1**. Twenty-six patients (61.9%) underwent procedures performed by surgical specialties (general/trauma, orthopedic, thoracic, vascular, urologic, or neurosurgical), and 16 (38.1%) underwent procedures performed by nonsurgical procedural specialties (gastroenterology, interventional radiology/neuroradiology, or cardiology).

**Table 1.** Baseline and procedural characteristics of patients with preexisting code-status limitations who died within 3 days of their procedure.

| Characteristic | n (%) |
| --- | --- |
| Age, median [IQR], y | 76 [66, 88] |
| Sex |  |
| Female | 16 (38) |
| Male | 26 (62) |
| Race |  |
| Black | 3 (7.1) |
| White | 34 (81) |
| Other<sup>a</sup> | 2 (4.8) |
| Unknown/Unavailable | 3 (7.1) |
| ASA Physical Status<sup>b</sup> |  |
| Low | 18 (43) |
| High | 24 (57) |
| Operative Stress Score<sup>c</sup> |  |
| Low | 31 (74) |
| High | 11 (26) |
| Periprocedural code status |  |
| Full code | 27 (64) |
| Not full code<sup>d</sup> | 15 (36) |
| Baseline serious illness category<sup>e</sup> |  |
| Cardiovascular | 22 (52) |
| Malignancy | 8 (19) |
| Neurologic | 6 (14) |
| Pulmonary | 5 (12) |
| Cirrhosis/liver disease | 3 (7.1) |
| None present | 4 (9.5) |
| Presentation |  |
| Related to chronic illness | 18 (43) |
| Injury or burn | 7 (17) |
| Infection | 7 (17) |
| Acute intraabdominal pathology | 4 (9.5) |
| Stroke | 3 (7.1) |
| Complication of treatment | 3 (7.1) |
| Surgical Service |  |
| General/trauma surgery | 10 (24) |
| Orthopedic surgery | 5 (12) |
| Thoracic surgery | 5 (12) |
| Vascular surgery | 3 (7.1) |
| Urology | 2 (4.8) |
| Neurosurgery | 1 (2.4) |
| Procedural Service |  |
| Interventional radiology/neuroradiology | 8 (19) |
| Gastroenterology | 6 (14) |
| Cardiology | 2 (4.8) |
| Procedure Urgency |  |
| Emergent | 16 (38) |
| Urgent | 4 (9.5) |
| Non-emergent | 22 (52) |
| Indication for procedure<sup>e</sup> |  |
| Treatment | 29 (69) |
| Palliation | 10 (24) |
| Diagnosis | 7 (17) |
| Life- or limb-saving | 3 (7.1) |

Abbreviations: ASA, American Society of Anesthesiologists; IQR, interquartile range.
<sup>a</sup> Asian, American Indian or Alaskan Native
<sup>b</sup> ASA Low = 2 or 3; High = 4 or 5
<sup>c</sup> Operative Stress Score Low = 1 or 2; High = 3, 4, or 5
<sup>d</sup> All patients presented for their procedure with a non-full code status. Periprocedural code status refers to the decision regarding code status intraoperatively and in the immediate post-procedural period. Maintenance of any limitation on resuscitation in the periprocedural period was defined as “Not full code.”
<sup>e</sup> Baseline serious illness category and indication for procedure were not mutually exclusive; percentages may sum to more than 100%.

The analysis produced a typology of four recurrent periprocedural terminal trajectory families: established terminal trajectory, abrupt terminal event, comfort-focused redirection after reassessment, and sustained rescue attempt. **Table 2** summarizes the defining and associated features of each family and presents its illustrative composite sequence. **Figure 2** shows the distribution of individual cases and key clinical and decisional elements across the periprocedural course; **Figure 3** presents schematic representations of each family’s recurrent sequence structure.

**Figure 2.**
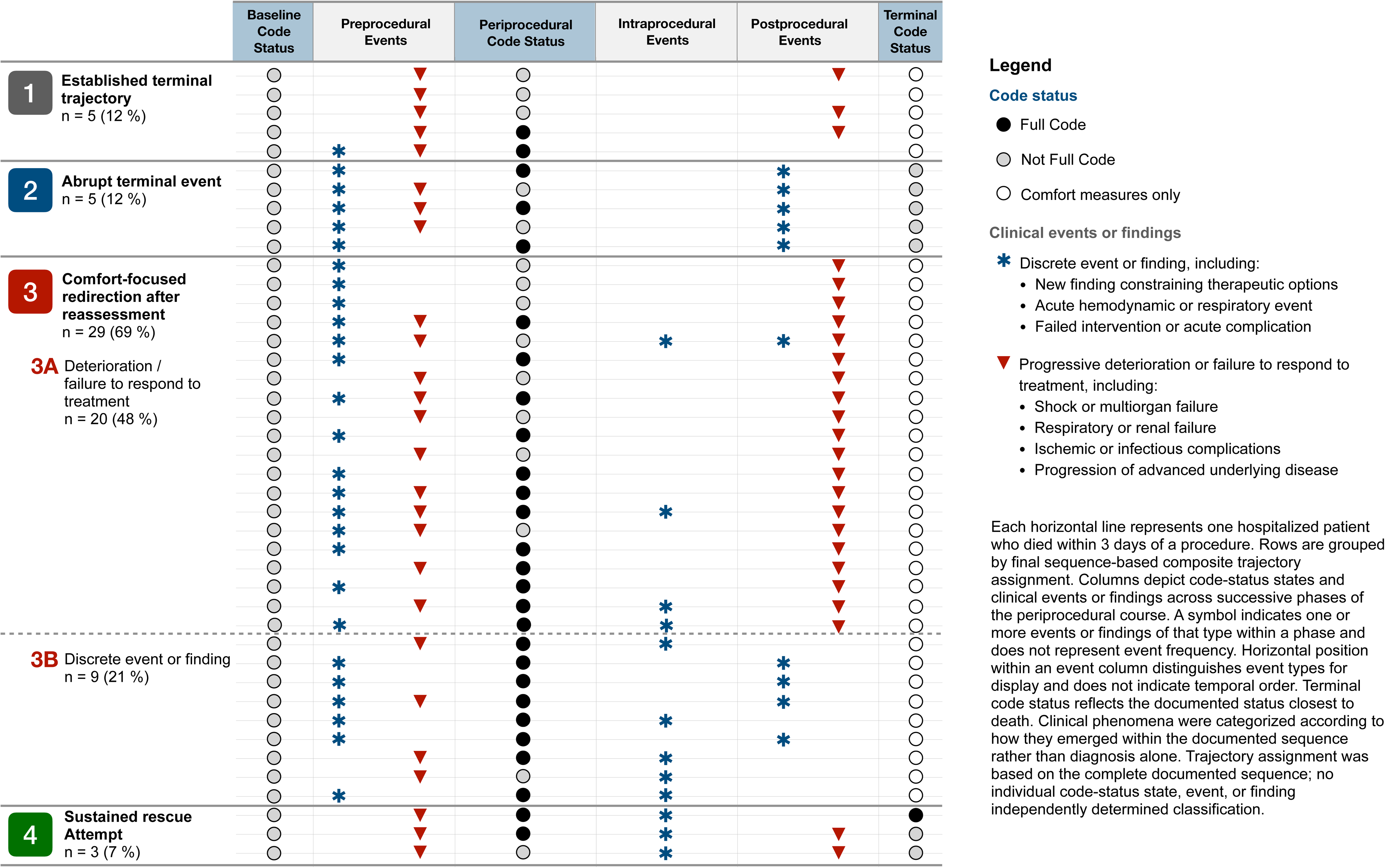
Clinical events and code-status transitions across terminal trajectories.

**Figure 3.**
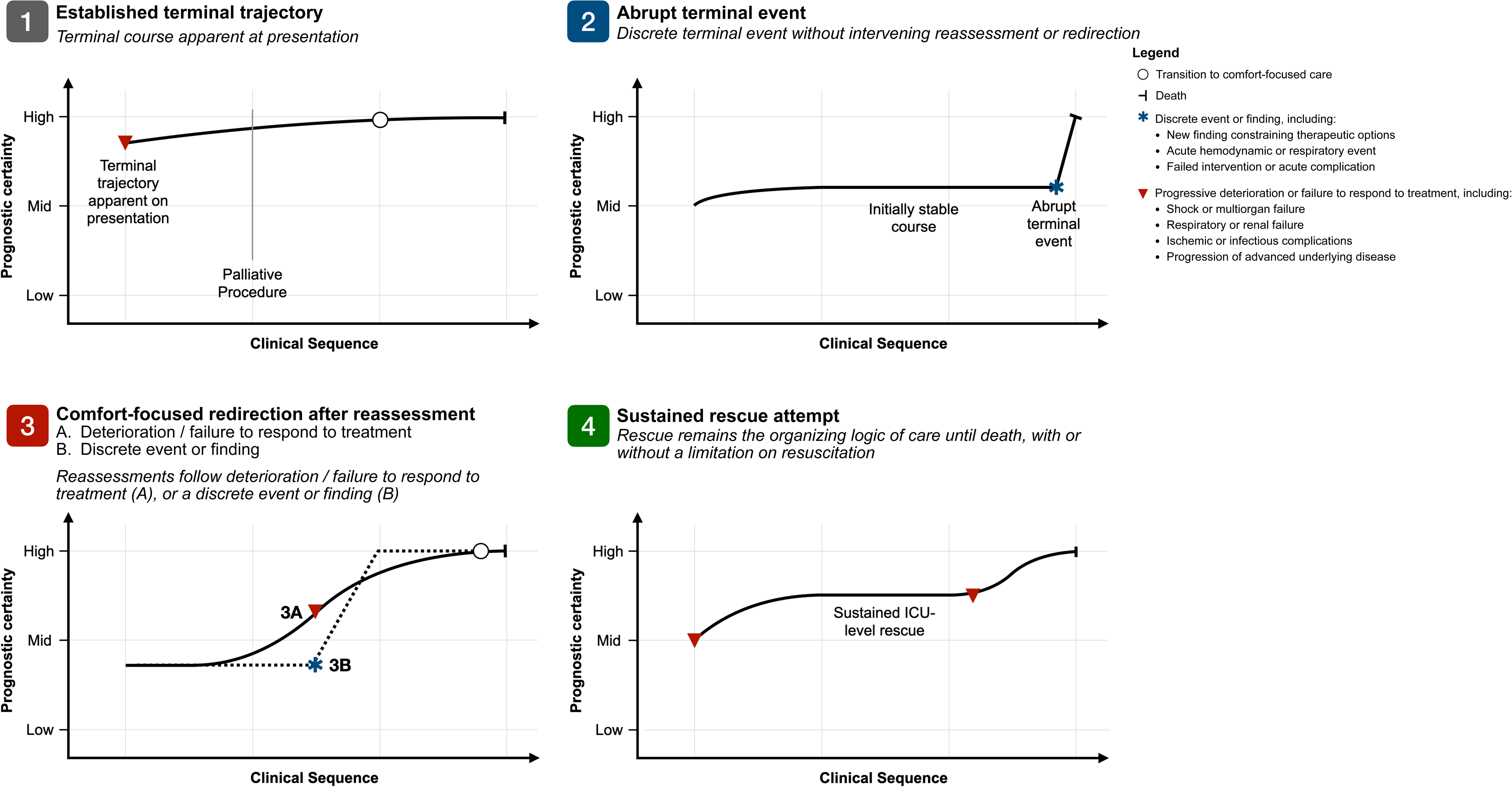
Schematic archetypes of clinical and decisional trajectories preceding early postprocedural death. The figure depicts recurrent clinical and decisional trajectory groups identified through qualitative sequence-based composite analysis. Curves represent clinical and decisional patterns synthesized across cases rather than quantitative estimates, patient-level measurements, or averaged trajectories. The horizontal axis indicates relative progression through the clinical sequence rather than measured time. The vertical axis represents relative prognostic certainty, defined as the degree of confidence regarding the likely clinical course as events and findings unfold. In the comfort-focused-redirection-after-reassessment panel, the solid trajectory depicts prognostic certainty emerging cumulatively through progressive deterioration or failure to respond to treatment, whereas the dotted trajectory depicts an abrupt increase following a discrete event or finding. The procedure is shown explicitly only for established terminal trajectories because its palliative intent and occurrence within a preexisting terminal framework are defining features of that trajectory; in the other trajectories, procedural timing and intent are contextual rather than category-defining.

**Table 2.**
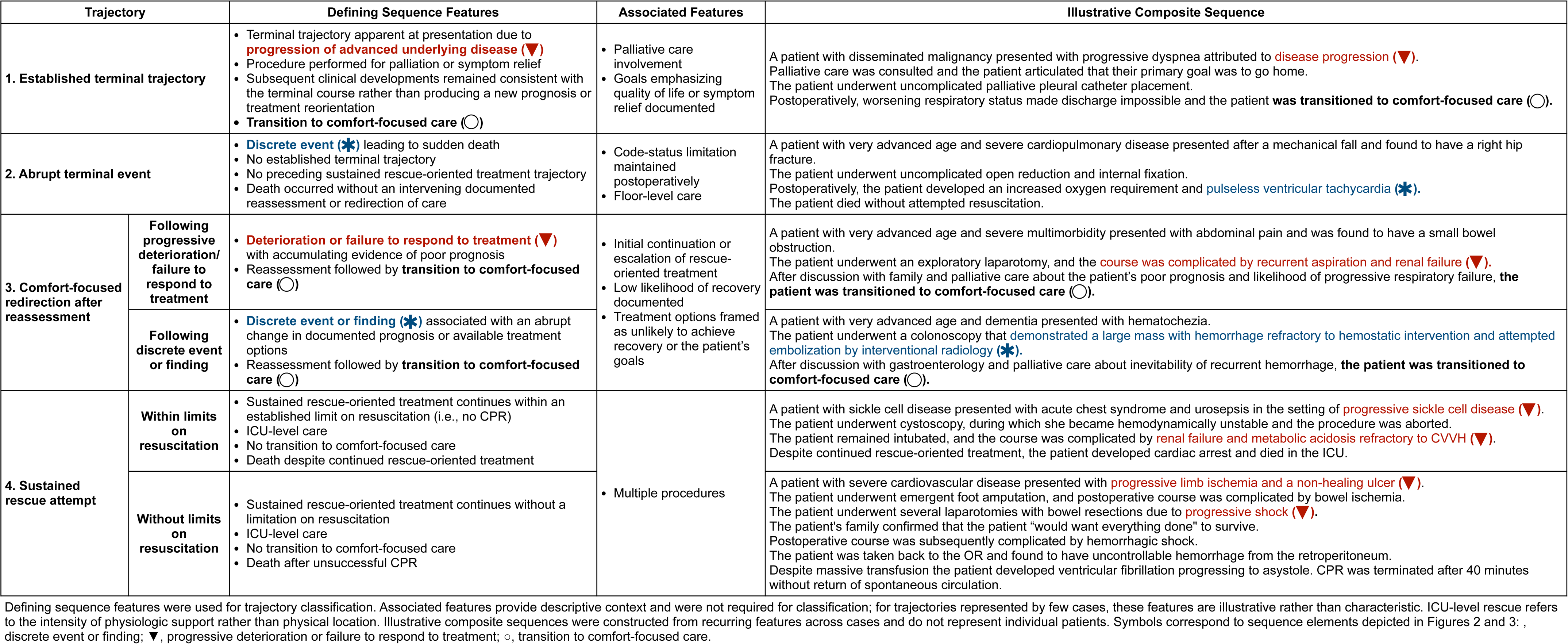
Terminal periprocedural trajectory typology.

### 1. Established Terminal Trajectory

Established terminal trajectory accounted for 5 deaths (12%). Patients entered the perioperative period with advanced terminal illness, and procedures were undertaken for palliation or symptom relief rather than treatment of the underlying disease process: 4 patients underwent pleural catheter placement and 1 received a venting gastrostomy tube. Subsequent clinical developments remained consistent with the terminal course rather than producing a new prognosis or treatment reorientation.

### 2. Abrupt Terminal Event

Abrupt terminal event accounted for 5 deaths (12%). These patients had neither an established terminal trajectory at presentation nor a preceding sustained rescue-oriented course. Sudden cardiac arrest in the postprocedural period led directly to death without an intervening redirection of care. Procedures included cardioversion, feeding tube placement, cystoscopy, and hip fracture repair.

### 3. Comfort-Focused Redirection after Reassessment

Comfort-focused redirection after reassessment was the most common trajectory family, accounting for 29 deaths (69%). In these cases, clinical developments were followed by documented reassessment of prognosis, available treatment options, or treatment goals and a subsequent transition to comfort-focused care. Two subtypes were identified: redirection after progressive deterioration or failure to respond to treatment (3A) and redirection after a discrete event or finding (3B). These patterns characterized how new prognostic information emerged rather than whether a procedural or treatment-related complication occurred; a complication could contribute to progressive deterioration or serve as the discrete event prompting reassessment and redirection.

Subtype 3A, redirection after progressive deterioration or failure to respond to treatment, accounted for 20 deaths (48%). The most common procedural subgroups were interventional radiology, open abdominal surgery, and endoscopy. The defining feature was persistent or worsening critical illness despite ongoing treatment, with accumulating evidence of poor prognosis rather than a single discrete event or finding. Treatment frequently continued or escalated during this course; documented reassessment was followed by transition to comfort-focused care.

Subtype 3B, redirection after a discrete event or finding, accounted for 9 deaths (21%). Events and findings were heterogeneous and included severe intraoperative instability, major aspiration with respiratory failure, failed interventions, and new pathologic findings associated with poor prognosis or limited treatment options. Despite their clinical heterogeneity, these events shared an abrupt change in the documented prognosis or available treatment options. Documented reassessment was subsequently followed by transition to comfort-focused care.

### 4. Sustained Rescue Attempt

Sustained rescue attempt accounted for 3 deaths (7%). In these trajectories, rescue-oriented treatment remained the organizing approach through death, without a transition to comfort-focused care. In 2 cases, intensive rescue continued within an established limitation on CPR, including mechanical ventilation, hemodialysis, and escalating vasopressor support. In the third case, rescue continued without a limitation on CPR and included repeated procedures, massive transfusion, and intraoperative CPR that was ultimately terminated without return of spontaneous circulation.

#### Reproducibility Assessment

The independent reviewer assigned 39 of 42 cases to the same trajectory family as the original consensus classification, corresponding to 92.9% overall agreement. Unweighted Cohen κ was 0.86 (95% CI, 0.70–1.00). All three disagreements involved cases initially assigned to comfort-focused redirection after reassessment. Post hoc review led to reclassification of one case as an abrupt terminal event in the final typology and minor clarifications to the composite presentations; the other two consensus assignments were retained. The reproducibility estimates remained based on the original consensus assignments.

## DISCUSSION

In this qualitative trajectory analysis of patients with preexisting code-status limitations who died within 3 days of a procedure, we identified four recurrent trajectories: established terminal trajectory, abrupt terminal event, comfort-focused redirection after reassessment, and sustained rescue attempt. Although all patients experienced the same outcome, their deaths arose through distinct relationships among baseline illness, procedural intent, clinical developments, prognostic reassessment, rescue-oriented treatment, and changes in treatment focus. The meaning of periprocedural code-status decisions and early mortality therefore depended on the broader clinical and decisional trajectory in which they occurred. Together, these trajectories provide an empirical framework for interpreting early postprocedural deaths among patients with treatment limitations. By identifying recurrent points at which prognosis and treatment direction changed, this framework can inform efforts to improve perioperative serious illness care through prospective treatment planning, communication about the intended scope of postprocedural rescue, and reassessment as the clinical course evolves.

Code-status documentation defines limits on specific interventions but captures only one dimension of periprocedural treatment decisions. In this study, preexisting limitations on resuscitative interventions coexisted with continued or escalated rescue-oriented treatment, whereas transition to comfort-focused care reflected a broader reorientation of the treatment plan. In the most common trajectory, treatment continued or escalated before reassessment and redirection; in sustained-rescue trajectories, it continued through death. Code status therefore did not map directly onto treatment intensity or overall therapeutic focus.

Among trajectories characterized by a transition to comfort-focused care following reassessment, we observed two broad patterns of new prognostic information: progressive deterioration or failure to respond to treatment, or a discrete event or finding that altered the documented prognosis or available treatment options. These patterns indicate that changes in treatment focus were temporally related to evolving prognostic understanding rather than fixed consequences of a preexisting code-status limitation. Prior work has identified preoperative expectations of commitment to postoperative life support,^12^ a broader default toward continued escalation of life-prolonging treatment,^13^ and clinical momentum generated by systems forces that promote surgical intervention and subsequent treatment cascades.^14^ Against this background, comfort-focused redirection represented an active change in therapeutic direction. Remer and colleagues recently demonstrated that new perioperative DNR orders cluster early after surgery and near death and are temporally associated with palliative care consultation.^15^ Our findings extend this work by reconstructing the clinical developments and documented reassessments that underlie documented changes in treatment limitations.

Viewed through this typology, “failure to rescue” is an incomplete explanation of early postprocedural mortality in this population. Failure to rescue identifies death following a complication, but it does not establish whether the complication was preventable or whether rescue was absent, unsuccessful, delayed, or no longer pursued after reassessment.^4,5^ Olson and Schwarze posed the more fundamental question of whether a decision not to pursue further life-prolonging treatment necessarily constitutes failure.^5^ Our findings show why that question cannot be answered from mortality or code status alone. The relevant question is not simply whether rescue culminated in survival, but whether continued treatment offered a reasonable prospect of achieving an outcome the patient considered worth its burdens.

Several comfort-focused redirection trajectories resembled time-limited trials: a defined period of life-sustaining treatment followed by reassessment and a decision to continue recovery-focused care, transition to comfort-focused care, or extend the trial.^16,17^ This framework has been proposed for older adults with frailty considering surgery when potential benefits and postoperative burdens are uncertain.^18,19^ Although not prospectively structured as time-limited trials, many trajectories followed similar logic: treatment continued while the clinical course generated new prognostic information, followed by reassessment of likely outcomes relative to the anticipated burdens and goals of further treatment.

These findings support prospectively framing selected high-risk procedures as one component of a time-limited trial. Under this framing, the procedure and subsequent treatments are not wholly separate decisions but phases of a single therapeutic course that warrants reassessment as the trajectory unfolds. Making this structure explicit could help teams define what the procedure is intended to accomplish and elicit which burdens of postprocedural treatment the patient considers acceptable.^20,21^ It could also help identify clinical developments that should prompt reconsideration of the treatment plan. This approach is consistent with efforts to operationalize “what matters” in surgical care by clarifying the outcomes treatment can realistically achieve, describing the burdens required to pursue them, and determining whether the patient considers those outcomes worth those burdens.^22,23^

Existing geriatric surgery standards recognize the need to revisit goals of care as the clinical course evolves. Current ACS Geriatric Surgery Verification standards require treatment goals to be updated before major procedures and upon significant changes in clinical status.^24^ Our findings provide specificity to these reassessment points by identifying trajectory-informed triggers, including progressive deterioration or failure to respond to treatment, a discrete event or finding that materially changes prognosis or available treatment options, and consideration of an additional procedure or life-sustaining therapy. These developments offer clinically meaningful points at which teams can reconsider with patients and surrogates whether possible treatments remain acceptable relative to their expected burdens and outcomes.

Several limitations warrant consideration. First, trajectories were reconstructed from clinical documentation and therefore could not capture conversations, rationales, or deliberations that were not entered in the EHR. Informal reassessments and decisions that did not result in a change in code status or treatment plan may therefore have been underrepresented. Second, this study was conducted within a single health system and intentionally focused on hospitalized patients with preexisting code-status limitations who died within 3 days of a procedure. The resulting typology characterizes early terminal trajectories in this selected population; it does not estimate the frequency of these trajectories among all patients with code-status limitations and may not apply to survivors, later postprocedural deaths, patients who died after discharge, or patients without documented preprocedural treatment limitations. Finally, this analysis was not designed to determine whether complications or deaths were preventable, whether care was goal-concordant, whether treatment limitations contributed causally to death, or whether different communication or decision-making strategies would have changed trajectories or outcomes.

In conclusion, early postprocedural death among patients with preexisting code-status limitations was not a uniform phenomenon but followed several distinct clinical and decisional trajectories. Mortality and code status alone could not establish whether rescue was constrained, sustained but unsuccessful, or redirected after new prognostic information emerged. Interpreting perioperative mortality and planning treatment therefore require attention to procedural intent, evolving prognosis, treatment, and reassessment across the broader therapeutic course.

## Data Availability

Data will not be made accessible to the public.

## Acknowledgments

Matthew Allen had full access to all the data in the study and takes responsibility for the integrity of the data and the accuracy of the data analysis.

